# Feasibility and Acceptability Study of Early Intervention Music Therapy for Young Children with Selective Mutism

**DOI:** 10.1101/2025.10.27.25338907

**Authors:** Kate Jones, Steve Gillard

## Abstract

**Introduction:** Selective Mutism (SM) is an anxiety disorder affecting some children starting school. Usual presentation is lack of speech at school, contrasting with confident speech at home. Estimated prevalence is 0.7%, rising to 2.2% for children who are second language learners. Long-term impacts include complex mental health conditions for children not receiving timely intervention. There are calls for effective treatments as part of a multi-modal care pathway. Case study research describes music therapy leading to generalised speech, but an evidenced-based approach is currently lacking. This study evaluates acceptability and feasibility of a manualised music therapy intervention for children with SM.

**Methods:** Four children with SM in early years school settings were offered manualised music therapy over a 14-week period. In-depth qualitative interviews were conducted with the music therapist, a school staff member and a parent of each child (n=12). Interviews were analysed using a framework grounded in well-theorised constructs of acceptability and feasibility.

**Results:** Acceptability was high for all participant groups, with clear comprehension and confidence in an intervention viewed as appropriate for the early years population. Need for cultural competence in delivery was highlighted. Analysis of feasibility demonstrated rapid recruitment, practicality and fit with the school environment, and a strong sense of perceived effectiveness including anxiety reduction, improved emotional well-being, increased self-expression and generalised speech.

**Discussion:** This study has added to the methodological literature on evaluating acceptability and feasibility in music therapy interventions and has indicated the appropriateness of moving to experimental testing of the SM intervention evaluated here.

## Introduction

Selective Mutism (SM) is an anxiety condition preventing people speaking in specific locations (e.g. school, social situations) despite normal speech in others (home) (APA, 2013). SM commonly presents when children enter the school system and is identified by the use of speech at home but not at school. Anxiety about speaking in specific settings is understood to be the most prominent causal factor in the development of SM (Anstendig, 1999; Cohan et al., 2008). Other factors such as a family history of shyness or SM (Steinhausen & Adamek 1997), parental mental health conditions (Koskela et al. 2020), social and general anxiety, childhood communication issues, developmental delay (Kristensen, 1997; Manassis et al., 2003), autistic features and behavioural inhibition (Muris et al., 2021) contribute to the current conceptualisation of SM having a multifactorial aetiology (Cohan et al., 2006; Muris et al., 2021).

Prevalence of SM in early years is thought to be around 0.7% (Bergman et al., 2002) and three times higher, at 2.2%, in children with immigrant backgrounds (Elizur and Perednik 2003). Evidence suggests that high levels of anxiety as well as parental orientation to mainstream culture, rather than bilingual status, explain increased vulnerability to SM (Starke 2018), although the nature of the association between SM, and cultural and linguistic diversity remains unclear (Slobodin 2023).

Awareness of SM is often poor (Kovac & Furr, 2019), with anxiety conditions prevailing into adulthood if SM is untreated (Koskela et al. 2023). Early detection (Schwartz et al., 2006) and intervention are a priority (Busse & Downey, 2011; Hung et al., 2012) to alleviate symptoms and prevent other mental health conditions evolving (Cline & Baldwin, 2004; Forrester & Sutton, 2015; Koskela et al 2023).

### Treatment for selective mutism

Behavioural interventions for SM view failure to speak as a way of avoiding anxiety connected with speaking. In their review, Cohan and colleagues (2006) concluded that behavioural techniques were successful interventions for SM, this continuing to be the dominant approach (Howe & Barnett, 2013; Jacob et al., 2013). Cognitive behavioural therapy (CBT), employing techniques such as psychoeducation, relaxation techniques, exposure and cognitive restructuring, has been successfully used to treat SM (Fung et al., 2002; Reuther et al., 2011; Ooi et al., 2012).

Involving parents in their children’s treatment has evolved as a key ingredient in some behavioural strategies. Ale and colleagues (2013) describe two case studies using systematic graduated exposure, with parents delivering exposure tasks. In an investigation of a conjoint behavioural-consultation framework, involving parents as active participants, Mitchell and Kratchowill (2013) concluded that a manualised approach would further assist in supporting school-based approaches for SM.

There are notable variations in the way behavioural approaches are delivered. Intensive SM ‘bootcamps’ in the US (Selective Mutism Center, n.d., Klein et al., 2017), and online CBT in Australia (Spence et al., 2008) address some of the practical, geographical demands of travelling to therapy sessions.

Selective Serotonin Reuptake Inhibitors (SSRIs) prescribed for a range of mental health conditions have also been used to treat SM, although evidence of effectiveness is limited due to small sample sizes and a lack of randomized controlled studies (Ostergaard 2018), while concerns are expressed by parents about the side effects for young children (Manassis et al 2016).

### Arts therapies

Arts therapies contribute to the treatment of SM, including dramatherapy (Oon, 2010), expressive play and art therapy (Fernandez et al., 2014), and arts therapies combined with behavioural techniques (Powell & Dalley, 1995; Moldan, 2005; Jackson et al., 2005). Cline and Baldwin (2004) state that music therapy ‘could have much to offer’ to the treatment of SM, but note the literature is sparse. Case reports have indicated that dramatic vocalisation (Amir 2005), loud playing of instruments (Monti, 1985, Sekeles, 1996; Mahns, 2003) and use of oral instruments (Trondalen, 2001; Von Moreau, 2005) are key elements of the music therapy process when working with people with SM. However, there is a lack of research describing music therapy as an intervention for children with SM, and no experimental evidence for music therapy’s efficacy as a treatment.

### Intervention development

The revised MRC/ NIHR complex intervention framework (BMJ 2021) describes the development and testing of healthcare interventions that might have multiple components, involve multiple actors in their delivery, and where implementation might be complicated by delivery context. Intervention development is driven by the existing evidence base, and through iterative cycles of engaging with stakeholders to refine underlying theory and core intervention components. The framework recommends combined assessment of the feasibility and acceptability of the emerging intervention to inform further refinement, before moving to formal evaluation of effectiveness.

Feasibility has been defined as an overarching concept to explore aspects of an intervention or of trial design for which more information is required before progressing (Eldridge et al 2016). Feasibility trials are designed to test the parameters of trial design – such as recruitment and retention of participants - and are increasingly common in music therapy research (Carr et al 2017; Porter et al 2018). Feasibility research might also address questions relating to specific uncertainties in intervention design, including qualitative enquiry into stakeholders’ views on the acceptability or specification of the intervention (Bond et al 2023). A mixed methods feasibility study of a bedside music therapy intervention for hospitalised critically ill patients tested patients’ interest, receptivity and satisfaction with the intervention (Fallek et al 2019). In a review of existing feasibility frameworks, Gadke and colleagues (2021) propose ten feasibility domains for more comprehensively testing interventions; recruitment capability, data collection procedures, design procedures, social validity, practicality, integration into existing systems, adaptability, implementation, effectiveness, and generalisability.

The acceptability of music therapy interventions has been assessed in various ways. Ma and colleagues (2024) used retention of participants in a randomised controlled trial as a proxy for acceptability of the intervention, while Grocke and colleagues (2013) and Carr (2014) use association between planned frequency of music therapy sessions and total number of sessions attended. Thompson and colleagues (2022) explore the acceptability of a novel neurologic music therapy intervention in a bespoke observational study from the subjective perspective of patients, staff and relatives, eliciting scaled responses to questions about how helpful participants found music therapy.

In the wider applied health sciences literature, acceptability has been more subtly theorised. Sekhon and colleagues (2017) conducted a review of reviews of the acceptability literature to develop a theoretical framework for assessing acceptability in healthcare interventions comprising seven component constructs: affective attitude; burden; ethicality; intervention coherence; opportunity costs; perceived effectiveness; self-efficacy.

### Developing and evaluating a music therapy intervention for children with SM

The iterative development of a theoretical framework, designed to underpin a manualised music therapy intervention for children with SM, is described first in inductive single case study research (Jones 2012) and subsequently through engagement with the wider SM case study literature and multiple case study research (Jones and Odell-Miller 2022). The latter study reported preliminary indication of the acceptability of music therapy as a treatment for SM, and the feasibility of integrating music therapy into school settings. The geographical location of these three studies was an inner-London borough.

A blueprint of an intervention manual for music therapy with children with SM was produced by the first author, informed by the theoretical framework, and then revised through iterative rounds of stakeholder consultation comprising four stakeholder groups; parents of children with SM, music therapists, members of a national SM clinical excellence network, and speech and language therapists working in early years settings. Although the term ‘manual’ is used, this was intended as a ‘toolkit’ and guidelines for best practice to be adapted flexibly to the needs of each child. The resulting intervention manual is used in the study reported below.

### Aim

This study aims to systematically evaluate the acceptability and feasibility of a manualised music therapy intervention for young children in early years school settings.

## Methods

### Study design

This paper reports a theoretically informed acceptability and feasibility study of manualised music therapy treatment for young children with selective mutism, in an early years school setting, comprising in-depth qualitative interviews with music therapists, school staff and parents of children referred to music therapy.

### Intervention procedures

Manualised music therapy for SM was delivered to four children, by different music therapists in four schools in an inner London borough. Therapists were trained by the first author using a training programme developed as part of the manual. Training took place as an online group, lasted for five hours, and comprised psychoeducation on SM, diversity awareness (provided by a colleague with specialist expertise), and a systematic but flexible approach to clinical practice, illustrated using case studies.

Children with SM in nursery or reception classes were identified and referred by Special Educational Needs and Disabilities Coordinators (SENDCO) after a presentation to the boroughwide SENDCO network meeting. SM was confirmed through consultation between researcher and school staff. Informed consent was obtained after providing participant information sheets to all parents inviting their children’s participation in the study. Therapy took place in carefully selected rooms in the children’s schools and was designed to take place once a week, lasting 30-50 minutes, for up to 14 weeks. There was a high degree of flexibility in delivery accorded to each child as specified in the manual.

The 70-page manual is split into chapters describing: background research informing development of the manual; description of SM and how it is currently supported in the UK; theoretical approaches to understanding SM; a process of Music Therapy intervention describing the approach, awareness of Speech and Language Therapy techniques such as sliding-in and generalisation (Johnson and Wintgens 2016), referral and assessment processes, working with the school team, and then a description of how key ingredients such as musical self-expression, oral instruments and humour (Jones & Odell-Miller 2022) can be utilised through the beginning, middle and ending stages of a therapy process.

Group supervision for music therapists was provided by the first author every 2-4 weeks (higher frequency at beginning and end of treatment) to consider fidelity to the therapy process, problem solve and highlight relevant resources.

### Participants

Participants in the study were the music therapist, a member of the school staff team and a parent/ carer of the child receiving therapy in each of the four schools (total n=12).

Music therapists were recruited to the study through an advert to members of the British Association of Music Therapy’s local network. Therapists who had completed the training described above were selected to participate on the basis of logistics and availability to deliver the intervention. School staff were identified as being someone who had most knowledge of the child in school.

All participants were first given detailed information about the study and invited to give written, informed consent to participate. Ethical approval for the study was given by Guildhall School of Music and Drama research ethics committee, on 4^th^ March 2024, reference number REA-2324-JONES-01.

### Data collection

Data were collected by in-depth, semi-structured interviews. Interviews took place using a secure online video-conferencing platform, were recorded using the platform and transcribed verbatim using Microsoft Word software. All interviews explored delivery of the intervention; interviews with music therapists also explored experience of training, use of the manual and supervision. Interview schedules were designed to elicit information about the feasibility and acceptability of the intervention.

### Data analysis

Data were analysed thematically informed by the feasibility (Gadke et al 2021) and acceptability (Sekhon et al 2017) frameworks. A first level coding organised data to the broad domains of acceptability and feasibility. Data were then coded to the distinct domains within the acceptability and feasibility frameworks. The *Social Validity* domain within the feasibility framework (Gadke et al 2021) covers constructs that overlap with many of the acceptability domains and so was not explicitly used in the analysis.

The analysis reports the different components of the intervention - training, manual, supervision and delivery - from the perspectives of parents (PAR), music therapists (MT) and school staff (ST) and by school (numbered 1-4).

## Results

All the children who received the intervention were aged 5 and attending school in reception classes. The children received between 9 and 13 sessions of Music Therapy with an average of 11 (one child went on extended holiday during term). Three White female music therapists and one White male music therapist participated in the study. All therapists were experienced with one having a long career in music therapy. Two Black mothers, one Black father and one White mother were interviewed about their children’s participation. School staff were three SENDCOs and one teaching assistant; two were White, one Black and one Asian, all were female.

### Training

#### Feasibility

In terms of *practicality*, therapists agreed that the training was ‘pitched at the right level’ (MT2), for ‘the right amount of time’ and with ‘enough breaks’ (MT3). One therapist described it as “clear and well-paced” (MT1) and another said it was “easy to grasp without being too simplistic” (MT4).

#### Acceptability

*Affective attitude* appeared predominantly positive, with one therapist describing the training as “incredibly helpful” and “without the training there’s no way I would have known anything” (MT4):

> “…really helpful and just a great introduction because although I had some awareness of what selective mutism was nothing to the kind of extent of the detail that you talked about…and learning about the techniques of like sliding in the speech” (MT4)

Another therapist said they felt “skilled up in a different area of my work that I never really had thought about before. It’s a great…intervention for children working with selective mutism and makes a lot of sense” (MT2).

Aspects of *intervention coherence* and *self-efficacy* within the training were noted. The fit between Music Therapy and treatments for SM was reflected upon. One therapist said that it was “kind of reassuring that there are sort of natural features of music therapy in the way that we all practice that plug into those existing ideas” (MT2). They did not feel as though they were going to have to “pursue a completely new approach” but instead focus on:

> “…selective mutism and the theories behind that and the kind of the way it ties into speech and language therapy, keeping all of that in mind…all of the sort of like key concepts like generalisation, desensitisation and sliding in” (MT2)

*Ethicality* was highlighted when two therapists mentioned the importance of the diversity component of the training, commenting that the trainer “not only added ideas from his own practice but gave a culturally sensitive perspective which is often missed out” (MT1).

### Manual

#### Feasibility

Three out of four of the therapists used the manual extensively but discussed its *practicality* in different ways. One therapist described how they printed it out as it felt “overwhelming as a digital document” (MT2). They then read the whole manual before starting therapy sessions, adding that it was “laid out well” (MT2). One therapist used the manual as a weekly “refresher” (MT3) and another described how “each week I would read more and the relevant bits to me” (MT4).

Therapists commented on “useful clinical examples” (MT2) and that “case studies were good to read about” (MT4). Another reflected that “real concrete examples of activities would have been helpful” but that “actually the musical elements section I found helpful” (MT3).

Therapists valued detail on more novel practical aspects supporting *adaptability* such as “ramping up the amount of sessions” (MT3) and “flexibility around session times, regularity” (MT2).

#### Acceptability

One therapist found the manual “reassuring”, “just gave me the confidence” and “I found it really, really helpful” (MT3). Another felt that it was “really comprehensive” (MT4). One therapist felt that “a lot of it was covered in the training” (MT1).

Therapists described the manual as *coherent*, indicating their confidence in, and understanding of the approach, commenting that “it’s backed up by evidence” (MT4), another describing it as “an umbrella or code of expectations” (MT2):

> “Having that permission, if you like not just permission but expectation that this works differently, that there are concepts behind this and that you know aligned with speech and language therapy” (MT2)

The novel concepts described were acceptable to another therapist who suggested that “the therapeutic process section was the most helpful” and emphasised their understanding of the importance of “using humour and keeping it light-hearted” (MT3).

*Intervention coherence* was also highlighted by a therapist who indicated what, for them, might have been missing from the manual. They described an article on SM from a psychotherapeutic perspective, stating it was ‘informative’ and ‘highlights the complexity’ (MT1).

### Supervision

#### Feasibility

The group supervision format seemed to have numerous benefits. Sharing clinical issues was the main advantage with therapists reflecting that it was helpful to have “everyone’s insights” (MT3) and to “brainstorm together” (MT4). They also highlighted the increased importance of supervision due to the “isolation” of working in a “specialist subject” (MT4).

Some suggestions were made as to how the supervision process could have been improved with “maybe slightly more regularity at the beginning”, “a WhatsApp group might have been good for quick questions and responses” (MT3) and “maybe a one-to-one at the halfway point” (MT4).

The availability of the supervisor for both practical and emotional support was highlighted:

> “I always felt like I could reach out to [the supervisor], [they’ve] made that really, really clear” (MT3)

#### Acceptability

Therapists indicated positive *affective attitude* towards group supervision describing it as “invaluable” (MT1), “very, very helpful” (MT3) and having “played a huge role” (MT4). One therapist said that it was “valuable to connect with others in the same boat” (MT2) and another said that “being on the same timeline was really interesting” (MT4).

Supervision appeared to contribute to strong *intervention coherence* and a positive sense of *self-efficacy* for the therapists, describing the importance of “feeling contained” and “having your expertise and reassurance” (MT4). They reflected on the need for “validation and support” (MT2), especially on therapy that was “anxiety provoking for those with children that are struggling” (MT4).

### Delivery

There was a wealth of data exploring acceptability in intervention delivery, and it was helpful to use Sekhon’s (2017) constructs of prospective, concurrent and retrospective acceptability.

#### Prospective acceptability

The *affective attitude* of all staff towards the intervention was positive and enthusiastic with comments such as “I think it’s a really nice therapy to provide” (ST1), and “I thought it was a really good idea for her” (ST4), demonstrating a strong fit with values (*ethicality*).

The o*pportunity costs* to schools of supporting the intervention were felt to be worthwhile, with staff describing how Music Therapy might provide a valuable addition in schools where resources are limited:

> “I just didn’t want to lose the opportunity, so very appreciative” (ST3)

Potential emotional *burden* was considered with one staff member observing a parent being “really apprehensive about it all and was a bit on the fence” (ST1). In another school, staff were again enthusiastic but mentioned the child’s trepidation:

> “I could see it was quite anxiety inducing for her. But I was so excited” (ST2)

Parents readily consented to their child’s participation, but some described both enthusiasm and caution about the emotional *burden*:

> “I had no concerns. It’s just ‘cause…she’s very shy. Like with people that she doesn’t know. So, it was more concerning for me if she was uncomfortable going.” (PAR1)

Overall *affective attitude* among parents was positive:

> “I thought, OK…I think it will be beneficial to have this therapy…and the idea to also to have your voice being heard. Or to bloom, I think that’s beautiful.” (PAR2)

#### Concurrent acceptability

Staff noted a shift in some children’s *affective attitude* and *burden* between initial and later sessions:

> “There were…quite a few tears at the beginning, but after a few sessions she got to be really comfortable with [the therapist], she said [the therapist] was her second favorite teacher” (ST2)

Parents had similarly positive observations:

> “It’s like he’s always looking forward to it. He’s always dancing, you know, and the songs he learns, he comes home singing it to his sister” (PAR3)

However, one parent shared *affective* and *ethical* concerns:

> “I thought that it would have been handy if her mentor [therapist] had also been a woman or a woman of colour. Not that [the therapist] wouldn’t necessarily be able to do it because of who [they were] or but just the ability to connect with someone more immediately and have faster growth” (PAR2)

Music Therapists also reflected on *affective attitude* and *burden* for the children receiving the intervention. One therapist described the high levels of anxiety at the beginning of the first session making it feel quite “touch and go” (MT2) as to whether the child would be able to attend. They went on to contrast this with the end of the session:

> “But at the same time during that session, you know, there was quite a lot of loud singing by the end of it.” (MT2)

To alleviate some of this *burden* the child attended the first few sessions with their teaching assistant. In contrast, the same therapist also described how:

> “I think I perceived in her a level of kind of like, drive, like, ‘I want to do this.’” (MT2)

There was also a sense of conflicting emotions once the child started to speak:

> “She was on a kind of rollercoaster of feeling proud and brave and happy to be doing that and to have found her voice. But then I think also scared and apprehensive when it started happening.” (MT2)

For one parent the additional emotional *burden* of their child attending therapy could have been better supported by improved “aftercare” (PAR2) or additional meetings with the therapist.

The amount and frequency of sessions was discussed with staff and parents expressing that “definitely more sessions would have been better” (ST3) and “not every day but maybe three times a week” (PAR4) to embed speech or increase progress, However one parent said:

> “Don’t think she was overly keen on twice a week…it would have probably been better to have longer sessions” (PAR2)

*Intervention coherence* appeared strong with one therapist describing how they felt “reassured that natural features of Music Therapy are right for SM therapy” (MT2), demonstrating an understanding of how many skills required for working with children with SM are already present in usual Music Therapy training.

#### Retrospective acceptability

Parents reported retrospectively their own and their children’s positive attitudes to Music Therapy:

> “I asked her funnily enough and she said she loved music therapy…I’m a very happy mommy that you’ve been helping my daughter.” (PAR4)

Reflecting on the *ethicality* of the intervention, teaching staff commented:

> “It feels almost softer to parents, doesn’t it? Than like speech and language therapy like, I think that can be an intimidating…music therapy feels like, lighter and nicer” (ST1)

Parents’ observations positively reflected on *perceived effectiveness*:

> “Just seeing how she is with her friends and saying more words before she wasn’t, she was just standing with a teacher” (PAR1)

When asked directly if their children had benefitted one parent said “100%!” (PAR1) and another “I feel that it was helpful” (PAR3).

This was true despite the challenging nature of some of the work such as one parent having sensitive, attuned support to reduce pressure on their child to talk:

> “I learned a lot from it…Like [I’ve] maybe been pushing [them to talk]” (PAR3)

One teacher commented on how aspects of generalising speech might have been improved:

> “…definitely more sessions would have been better, and I think the other thing was and I know this was out of your control, but having a friend during the sessions and I think that was agreed like the day before the last session or something so” (ST3)

For this study, ethical approval for adding a friend to support generalisation was delayed although written into the manualised intervention, demonstrating how teachers’ perceptions of what might be effective wholly aligned with the intervention manual.

Enhanced *self-efficacy* was reflected upon by one therapist:

> “I feel skilled up in a different area of my work that I never really had thought about before. It’s a great, it’s a great Intervention for children working with selective mutism and makes a lot of sense” (MT4).

#### Feasibility

*Recruitment capability* was a key dimension to consider as part of the study. Potential referrals were taken rapidly from SENDCOs during and after the borough network meeting (10 in the first week):

> “I remember when you came to the SENDCO meeting and you talked about it, I was like, I think emailing you as you were talking” (ST1)

Some staff suggested practical improvements to the intervention:

> “…maybe catching it earlier…so maybe in the autumn term would have been, you know, not saying that this hasn’t been beneficial, but to have them from the beginning, so you can implement it straight away”. (ST4)

Another therapist’s comments suggested a strong link between fidelity to the manual (*implementation*) and *effectiveness* describing their “quite light and playful” approach and how the child was “very responsive to that straight away” (MT4).

*Integration* and the alignment of aspects of the intervention with school culture, such as teamwork, was commented on by school staff:

> “She sent me an email each week of just a little paragraph updating how the session went, and then I would then forward that to the mum because the mum was a little bit apprehensive to begin with about everything…. it’s just really quick and efficient…it’s just helpful for my own record keeping and evidence building” (ST1).

Staff comments suggested a good fit between Music Therapy and the school system generally:

> “…it would be so lovely for something similar to be able to be brought out for the other children in my class, it would be massively beneficial I think, we’ve been really lucky” (ST2).

More concrete, rather than perceived indicators of *effectiveness* are a critical element of feasibility, with the intervention designed to impact a range of therapeutic outcomes such as anxiety reduction, improved emotional well-being, increased self-expression and speech. Therapists’ comments reflect an understanding of this:

> “Yes, I noticed that from being completely silent, she started making a vocal sound, you know, quite close, something like a mix of discomfort and saying ‘no’ to something and then I think two sessions after that, she’s suddenly started using ‘yeah’, made other vocal sounds and then started to use the words all of a sudden” MT1

They also described how the child “became more relaxed. So that felt very nice” (MT1). Another therapist described progress with emotional wellbeing and speech:

> “She seemed quite down and gloomy before and very kind of isolated, but as the therapy went on, she was happy to come into school. And that she was putting herself forward and doing things that she wouldn’t have done before. And certainly I saw her every time I went to collect her. In the last five weeks or so, she was playing with other children, talking and chatting and doing lots of role play”. (MT4)

This generalisation of speech from therapy room to classroom is a key measure of *effectiveness*. During the study three of four children spoke in the therapy room, with all four children starting to speak within their school environment. One therapist was unaware of this speech progress in school reporting that they were “really surprised he’s not spoken” within the therapy sessions (MT3). The pupil’s teacher described their student’s outcomes - speech and improved emotional well-being - through an educational lens:

> “When he’s reading, he does say words. But the actual reading is really, really important. We’re very happy with that” (ST3)

Reading from the school report they also shared:

> “So he’s had an excellent year reception because every morning he arrives with a big smile and explores the classroom to see what he wants to take part in. He’s popular and friendly. Very positive approach to his learning.” (ST3)

Another child changed from being “frozen” (MT2) to being very socially confident:

> “She’s become really good friends with a few of the really confident children in the class.” (ST2)

The teacher also commented on their speech generalization:

> “So when we’ve done circle times recently, she’s much more confident to speak and a full sentence.” (ST2)

One school staff member reflected on the potential *generalisability* of skills from Music Therapy into the school environment and were keen for “nuggets” of skills sharing to embed that generalisation:

> “I think it would have been helpful to…have really concrete strategies that like could feed from like in this week’s session…especially being in reception where they have that continuous provision. So things are almost most easily embedded down there” (ST1)

## Discussion

This study demonstrates strong acceptability and feasibility of delivery of a well-theorised Music Therapy intervention for young children with SM in early years educational setting, developed following MRC complex intervention guidance (BMJ 2021).

### Acceptability

Our findings indicate that the training programme and manualised intervention is highly acceptable to Music Therapists across the seven constructs of Sekhon and colleagues’ (2017) framework. Therapists emphasised the importance of prior theoretical development work (Jones 2012; Jones & Odell-Miller 2022), engendering confidence in the intervention, and were reassured that usual features of Music Therapy practice provide essential ingredients that also align with Speech and Language Therapy interventions for SM (Johnson & Wintgens 2016). These high levels of comprehension and confidence enabled the intervention to be implemented as manualised. It is interesting to note that one therapist did share a psychotherapeutic approach to treatment of SM they thought might be helpful (Monzo et al. 2015). This source described SM as a ‘choice’ while citing many now outmoded theories, reinforcing the importance of accurate formulation of SM underpinning clinical work (Jones & Odell-Miller 2022). Therapists also indicated that group supervision provided a particularly successful and acceptable mechanism for supporting intervention delivery and fidelity.

The analysis told us that school staff view Music Therapy as an appropriate intervention for young children and as a result are overwhelmingly positive about referring children, demonstrating trust in the approach. This may have been in part due to local, historical factors such as prior positive experience of the Music Therapy service locally, as well as two previous Music Therapy and Selective Mutism studies with promising results occurring within the geographical location of this study (Jones 2012, Jones 2019). Staff also referred to a paucity of resources for supporting children’s specific needs within primary schools, highlighting that children with SM frequently don’t reach the threshold for an education and health care plan (EHCP). As such, a music therapy intervention for SM responds to calls in the new NHS 10-year plan to accelerate the roll-out to schools of evidence-based, early interventions from allied health professionals to address child mental health and special educational and disability (SEND) needs (DHSC 2025).

Our data indicated a willingness among parents to consent to, and positivity towards the prospect of their children attending Music Therapy sessions, maintained throughout the intervention. An additional consideration was that, at this pre-therapy stage, parents were being informed - some for the first time - that their children had SM. However, this did not appear to alter their positivity towards the intervention.

The study demonstrated the importance of the diversity awareness component of the training, highly valued by the therapists and underscored by a parent’s reflections on the potentially poor fit between the therapist (White) and their child (Black). The prevalence of children with SM increases three-fold in inner-city boroughs where levels of immigration are high (Elizur & Perednik 2003) indicating that cultural factors deserve further exploration in SM research (Slobodin 2023). Diversity of therapists and cultural awareness within sessions remains important work. UK Music Therapists are beginning to unpack and address the impact of colonial attitudes within training and therapy practices but sustained commitment to this work is needed (Mains et al 2025).

An important finding of the study was recognition of the sensitive consideration needed to assess the potential emotional burden of the intervention for children with high levels of anxiety. Staff and parents expressed concern that anxiety might impact initial attendance at sessions. In addition, the nature of SM frequently evokes a sense of urgency and pressure on children, parents and therapists to achieve speech production (Johnson & Wintgens 2016). Awareness of this and potential emotional fatigue must be factored into supervision and project management. One parent suggested that this aspect of the intervention might be better supported with increased ‘aftercare’. Children overwhelmingly engaged well and benefitted from the intervention although some became strongly attached to their therapists finding endings challenging. Overall, the results signal strong levels of acceptability despite high participant anxiety.

### Feasibility

Assessment of recruitment capability was a key dimension of the study. Gadke (2021) describes an SM study (Bergman 2013) that experienced recruitment challenges due to the low prevalence rate of SM. In our study, multiple referrals of appropriate participants were made in a short period of time following a presentation of the research t the SENDCO network locally. As noted above, this study was optimally located in a site of prior successful research (Jones 2012, Jones & Odell Miller 2022) within a long-standing Music Therapy service. Our findings suggest that replication of similar communication strategies with experienced researchers would strengthen recruitment capability in a larger study. In addition, no children or therapists dropped out of the study.

The study demonstrated the practicality of intervention delivery (Gadke 2021), across the one-day training, supervision and described therapy set-up. Staff commented that an earlier start in the academic year may have been beneficial. Most children and parents were happy to schedule twice weekly sessions later in therapy to embed progress reflecting flexibility as described in the manual. The intervention appeared to integrate well into the educational system, with some teachers expressing the wish for more Music Therapy for other children.

Perhaps the most noteworthy component in the analysis of the acceptability and feasibility of this intervention is that it was perceived to be effective - among teachers, parents and therapists - for all the children across a range of different domains, including anxiety reduction, improved emotional well-being, increased self-expression and speech. With SM, speech generalisation across the school environment is key (Johnson & Wintgens 2016) and this occurred with all four children, offering strong indication of likely effect and suggesting the readiness of the intervention for formal testing in a randomised clinical trial.

### Strengths and limitations

It is a strength of this study that it was conducted as part of a sequence of research, iteratively developing and refining the intervention, theoretically and empirically (Jones 2012; Jones & Odell-Miller 2022), as specified in the MRC complex intervention framework (BMJ 2021). The framework recommends concurrent testing of feasibility and acceptability, and this study did so systematically, grounding that assessment in clearly specified constructs (Gadke et al 2021; Sekhon et al 2017).

The number of cases (four) was small, limiting the range of potential issues that might have been explored. We did not have opportunity to assess the full range of flexibility components within the manual; for example, inviting peers into therapy sessions.

However, a comprehensive data set (parent, therapist and school staff) was collected for each case, enabling in-depth assessment from multiple perspectives across nearly the entirety of the manual. Children were not interviewed - this would have been methodologically and ethically challenging because of their age and condition – but it is noted that in-depth multiple case study work with young children had informed development work underpinning this study (Jones & Odell-Miller 2022).

Findings were qualitative and as a result we lacked concrete measures of some aspects of feasibility and acceptability (e.g. delivery of specific intervention components).

However, the qualitative approach provides considerable insight informing further refinement of the manual prior to delivery in a trial.

### Implications for music therapy research

We noted above that testing of feasibility and acceptability in music therapy research to date has been largely limited to a small number of studies assessing quantitative indicators of feasibility in pilot trials (Carr et al 2017; Porter et al 2018) or proxy indicators of acceptability (Grocke et al 2013; Carr 2014). In recent years, the music therapy profession has moved from a focus on evidence-based practice, to developing and delivering evidence-based interventions (Aigen 2015), with work still to be done in the reporting of high-quality intervention research and in particular sufficiently detailed specification of interventions (Robb et al 2018, 2025). This study offers a template for systematic development work to inform delivery and reporting of high-quality randomised controlled trials of music therapy interventions.

## Conclusion

This study indicates that Music Therapy offers an acceptable and feasible intervention for young children with SM in early years educational settings. The study also informs further refinement of the intervention prior to formal testing of efficacy in a randomised controlled trial.

## Data Availability

All data produced in the present study are available upon reasonable request to the authors

## Acknowledgements

Alphonso Archer contributed to development and delivery of cultural competence aspects of the intervention training. Anita McKiernan advised on development of the intervention from a highly specialist clinical perspective.

